# COVID-19 mortality in children and young people in Mexico

**DOI:** 10.1101/2021.09.01.21262981

**Authors:** Dalia Stern, Eduardo Arias de la Garza, María Teresa García-Romero, Martin Lajous

## Abstract

**Objective:** To estimate COVID-19 and pre-pandemic low respiratory infection (LRI) mortality in children and young people in Mexico.

**Material and methods:** We estimated the percentage of total mortality attributable to COVID-19 (95% confidence intervals; 95%CI) and corresponding estimates for pre-pandemic LRI mortality.

**Results:** In 2019, LRIs represented 8.6% (95%CI 8.3, 8.9) of deaths in children 0-9 years and 2.0% (95%CI 1.8, 2.3) in those aged 10-19 years. In 2020, corresponding estimates for COVID-19 were 4.4% (95%CI 4.1, 4.6) and 3.7% (95%CI 3.4, 4.1).

**Conclusions:** Relative to LRI, COVID-19 may be exerting considerable mortality burden, particularly in older children and adolescents.

## INTRODUCTION

In high-income countries (HIC), COVID-19 deaths in children and young people appear to be uncommon.(1) However, in low- and middle-income countries pediatric care may be less resilient to the challenges faced by the current pandemic.(2) In Mexico, even while adolescents and young adults may have been particularly important for SARS-Cov-2 transmission,(3) the observed number of deaths in persons ≤20 years of age remains lower than expected.(4) Also, while COVID-19 may be within the first ten causes of death in this age group, in 2020 deaths due to perinatal and congenital conditions, malignancies, and injuries may have exerted a higher burden.(5) As school activities resume and vaccination efforts advance, understanding the patterns of mortality within this age group before and during the pandemic may provide important context to parents, clinicians, and decision makers. Thus, we estimated COVID-19 and pre-pandemic low respiratory infection (LRI) mortality in children and young people in Mexico.

## METHODS

We obtained death certificate-based all-cause and COVID-19 mortality for people aged <1, 1-4, 5-9, 10-14, and 15-19 years in Mexico between January 1^st^, 2020 and July 10^th^, 2021.(6) We also used epidemiologic surveillance data for PCR- or antigen-confirmed SARS-CoV-2 deaths (March 1^st^, 2020-July 10^th^, 2021).(7) We estimated age-specific cumulative all-cause, COVID-19, and lab-confirmed COVID-19 deaths per 100,000 for 2020 and for January 1^st^ 2021-July 10^th^ 2021 using mid-year population estimates.(8) We calculated percentage of total mortality attributable to COVID-19 and 95% confidence intervals (95%CI; shown in **Supplementary Tables 1 and 2**). For comparison, Global Burden of Disease 2019 all-cause and LRI deaths were used to calculate corresponding 2019 LRI estimates.(9)

### Results

Relative to 2019, 2020 all-cause deaths for children and young people were lower (37,279 vs. 44,439) and the distribution across age groups changed (e.g., % of all deaths in <1 year: 56.8% in 2019 and 44.6% in 2020; **Table 1**). In 2019, LRI represented 8.6% (95%CI 8.3, 8.9) of deaths in children 0-9 years and 2.0% (95%CI 1.8, 2.3) in those aged 10-19 years. In 2020, 4.4% (95%CI 4.1, 4.6) of deaths in children 0-9 years were attributable to COVID-19. In the 10–19-year group, the corresponding estimate was 3.7% (95%CI 3.4, 4.1). Adolescents and young adults aged 15-19 years appear to be driving the mortality rate in this group, however, the proportion of deaths attributable to COVID-19 in different age groups appears to be similar (10-14 years 4.0%, 15-17 years 3.6%, 15-19 years 3.7%, 18-19 years 3.7%, and 12-17 years 3.7%). Compared to 2020, between January 1^st^ and July 10^th^ 2021, we observed a lower proportion of deaths attributable to COVID-19 (2.1; 95%CI 1.7, 2.4) in children <1 year. For those aged 10-19 years, the proportion of deaths attributable to COVID-19 remained at 3.6% (95%CI 3.4, 4.1).

**Table 1.**
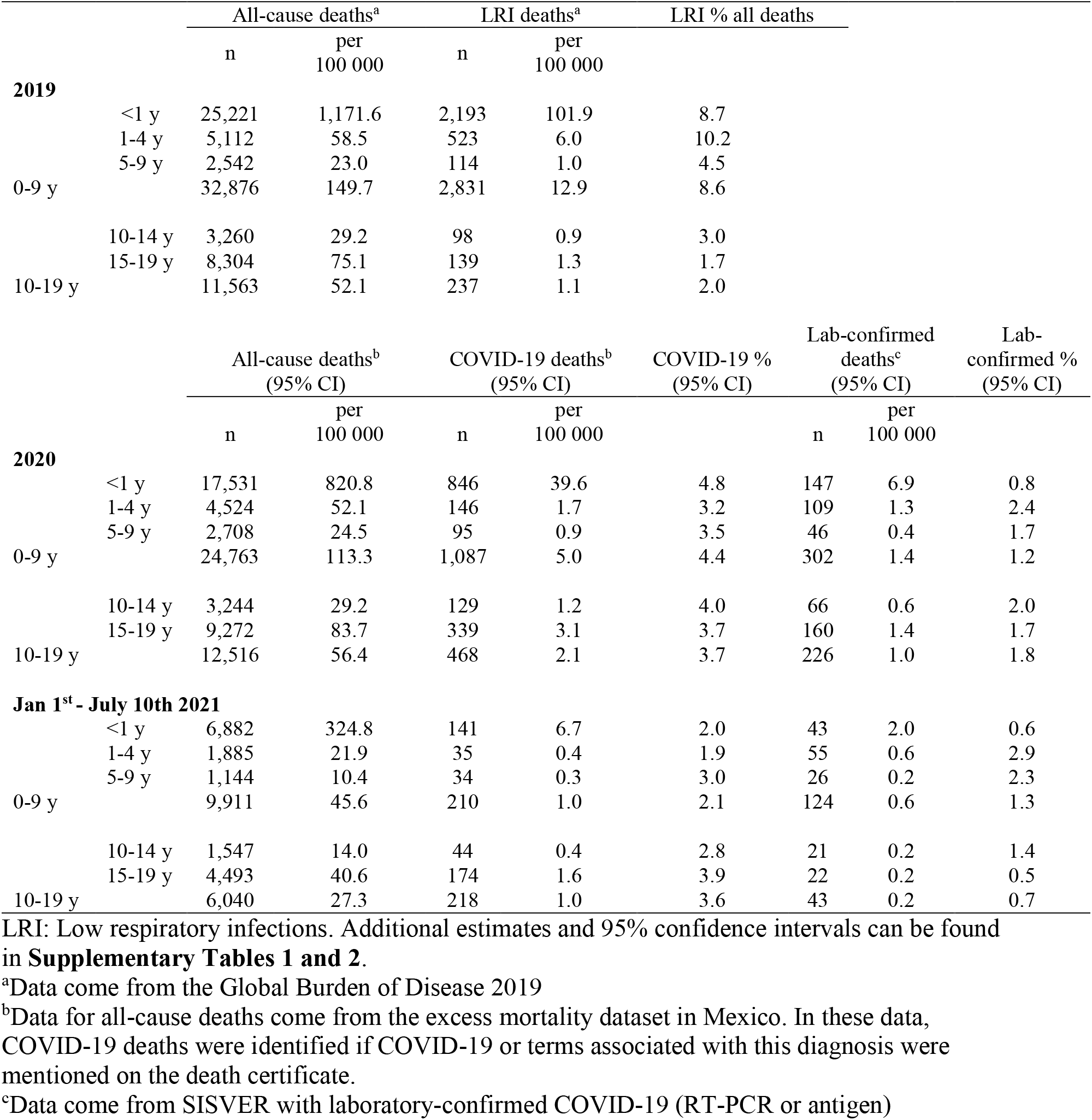
Age-specific data for Mexico showing all-cause and LRI mortality for 2019 and all-cause and COVID-19 mortality for 2020 and 2021.

Lab-confirmed COVID-19 deaths were substantially lower for all age groups. In children 0-9 years, 302 COVID-19 deaths (1.2% of all deaths; 95%CI 1.1, 1.4) were lab-confirmed and in the 10–19-year age-group 226 deaths were recorded (1.8% of all deaths; 95%CI 1.6, 2.0). There was an overall increase between 2020 and 2021 in lab-confirmed relative to COVID-19-certificate attributed deaths (34.0% to 39.0%). While in the <1-year age group, this proportion increased dramatically (17.4% in 2020 vs 30.5% in 2021), in those aged 15-19 years, this proportion decreased from 47.2% to 12.6%.

Between January 1^st^ 2020 and July 10^th^ 2021, the largest increase in cumulative COVID-19 mortality was observed in children <1 year and in those aged 15-19 years (**Figure 1**). At the first pandemic peak (July 16^th^ 2020), adult cumulative mortality was 57 versus 23 per 100,000 in children <1 year and 1.1 per 100,000 in those aged 15-19 years. By the second peak (January 18^th^ 2021), the corresponding estimates were 194 per 100,000 for adults, 41 per 100,000 for children <1 year, and 3.3 per 100,000 for adolescents and young people aged 15-19 years.

**Figure 1.**
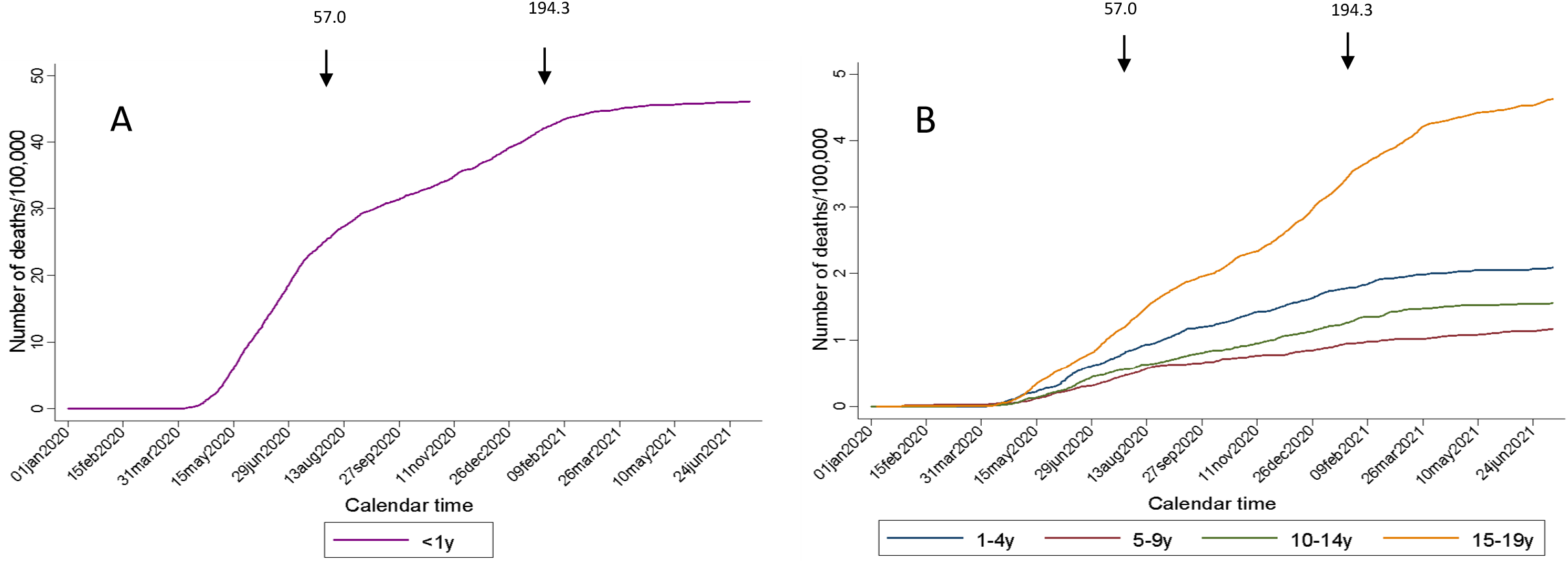
Trends of child and young person mortality from COVID-19 in Mexico between January 1^st^, 2020 and July 10^th^ 2021. A) <1 year; B) 1-4 years, 5-9 years, 10-14 years, and 15-19 years. Arrows denote the date of peak all-age reported mortality per day from COVID-19. Numbers above arrows correspond to the cumulative mortality/100,000 people on the date of peak for all-age groups in Mexico.

## Discussion

COVID-19 mortality in children and young people in Mexico was higher as compared to HIC. The mortality burden of COVID-19 among those aged 10-19 years may be higher than that of LRI before the pandemic. Yet, for younger children, COVID-19 mortality was lower than for pre-pandemic LRI. However, relative to adults and other causes, COVID-19 mortality in children and young people in Mexico has remained low.

LRI mortality in people<20 years is uncommon in HIC (2% of total deaths).(9) Mexico still lags behind other middle-income countries like Argentina (5%) and Chile (3%). Mexico’s LRI mortality in people <20 years (7%) may reflect poor healthcare access and substandard quality of care. Also, relative to HIC, Mexico’s COVID-19 mortality burden in this age group appears to be higher. In the United Kingdom by early 2021, COVID-19 mortality was 0.1 and 0.3 per 100,000 in children and young people aged 0-9 years and 10-19 years, respectively.(1) Based on death certificates, the corresponding estimates for Mexico were 5 and 2.1 per 100,000. Lab-confirmed COVID-19 mortality rates provided a lower bound of the mortality rate (1.4 and 1.0 per 100,00 for ages 0-9 and 10-19, respectively). Also, the proportion of deaths attributable to COVID-19 in 2020 was noticeably higher than in HIC (1) and was almost two times of what was observed for LRI prior to the pandemic in those aged 10-19 years (3.7% vs. 2.0% of all-cause deaths). Among children 0-9 years, 4.4% of all-cause deaths were attributable to COVID-19 (vs. 8.7% for LRIs). COVID-19 mortality and young people may echo health system challenges to provide adequate care for LRI in this age group.

Our findings may be limited by our reliance on error-prone death certificate information. For COVID-19 the magnitude of the error is possibly higher than for LRI, could be age group-dependent, and may have changed over time. In addition, our use of 2019 for comparison may seem arbitrary given yearly influenza epidemics. However, variation in LRI deaths was minimal in 2015-2019 in Mexico (7) and the fraction of LRI mortality attributable to influenza in children is below 2%.(10) We could not rely on lab-confirmed COVID-19 because SARS-Cov-2 testing has been limited in Mexico, particularly for children and adolescents. However, we presented results on lab-confirmed COVID-19 bound COVID-19 mortality estimates. COVID-19 deaths may include high-risk populations with co-morbidities (i.e., cancer), which could not be identified in this study. Finally, the mortality rates for the first half of 2021 are not comparable with those for 2020. Also, estimates for the first half of 2021 may be less reliable because of the increasing importance of SARS-Cov-2 variants of concern, the seasonality of respiratory infections, and reporting lag.

Relative to pre-pandemic LRI deaths, COVID-19 may be exerting considerable mortality burden, particularly in older children and adolescents. Our conclusion differs from a previous assessment that limited the comparison to COVID-19 mortality burden in adults and focused on Mexico City only.(11) While most SARS-CoV-2 infections in children and young people will likely result in mild disease,(12) community transmission will increase demand for specialized pediatric care. As vaccination efforts in adults advance in Mexico and information on the potential harms of vaccinating younger age groups emerge, policy makers should weigh the benefit of vaccinating older children and adolescents in a setting where pediatric care for LRI has been suboptimal and when COVID-19 has become a vaccine-preventable disease.

## Data Availability

All data are publicly available

---

**Supplemental Table 1:**
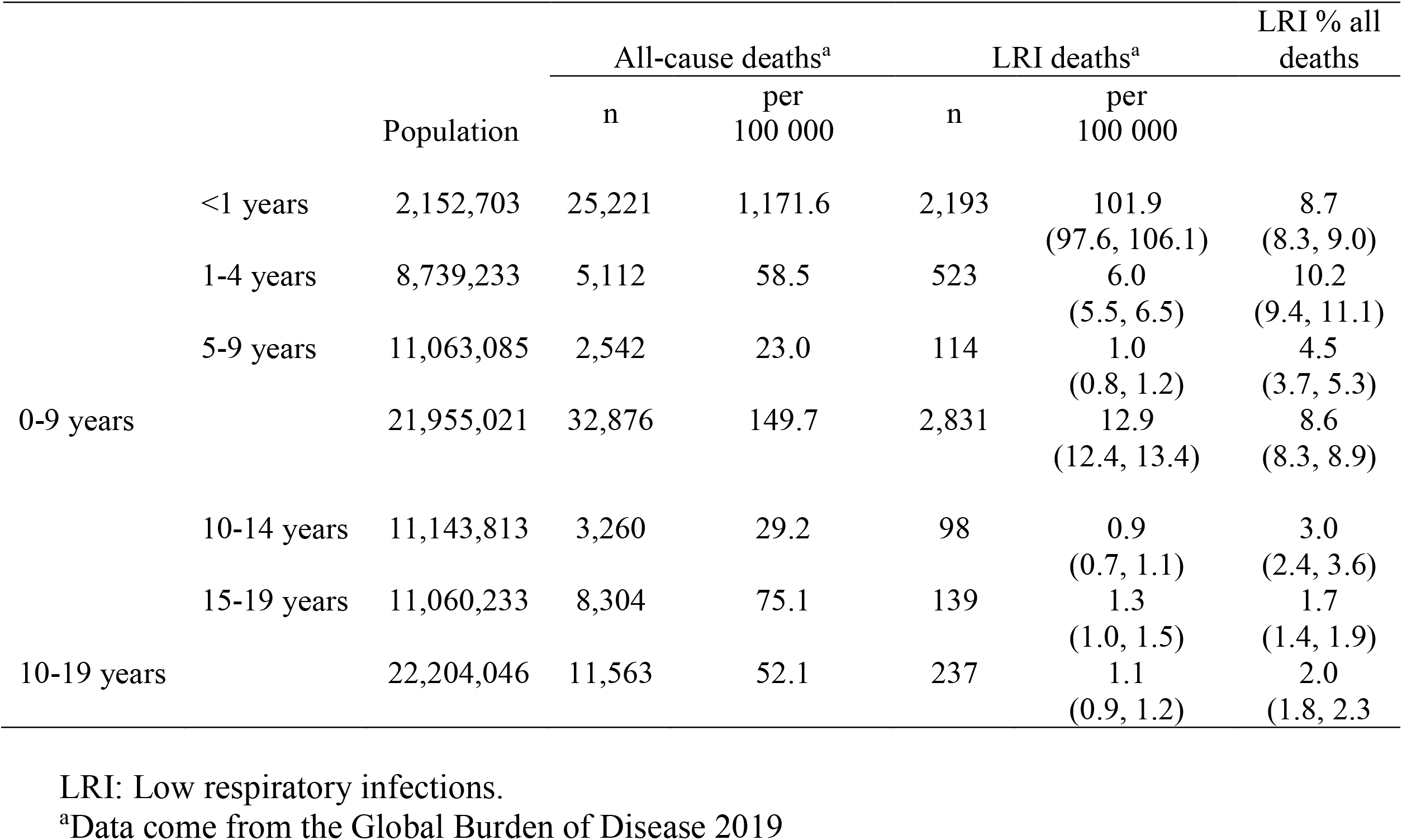
Age-specific data for Mexico showing all-cause and LRI mortality and 95% confidence intervals (95%CI) for 2019.

**Supplemental Table 2:** Age-specific data for Mexico showing all-cause and COVID-19 mortality for 2020 and 2021.

|  |  |  | All-cause deaths<sup>a</sup> |  | COVID-19 deaths<sup>a</sup><br>(95% CI) |  | COVID-19 %<br>(95% CI) | Lab-confirmed deaths<sup>b</sup><br>(95% CI) |  | Lab-confirmed %<br>(95% CI) |
| --- | --- | --- | --- | --- | --- | --- | --- | --- | --- | --- |
|  |  |  | n | per<br>100,000 | n | per<br>100,000 |  | n | per<br>100,000 |  |
| **2020** |  | Population |  |  |  |  |  |  |  |  |
|  | <1 years | 2,135,722 | 17,531 | 820.8 | 846 | 39.6<br>(36.9, 42.3) | 4.8<br>(4.5, 5.1) | 147 | 6.9<br>(5.8, 8.0) | 0.8<br>(0.7, 1.0) |
|  | 1-4 years | 8,678,435 | 4,524 | 52.1 | 146 | 1.7<br>(1.4, 2.0) | 3.2<br>(2.7, 3.7) | 109 | 1.3<br>(1.0, 1.5) | 2.4<br>(2.0, 2.9) |
|  | 5-9 years | 11,035,276 | 2,708 | 24.5 | 95 | 0.9<br>(0.7, 1.0) | 3.5<br>(2.8, 4.2) | 46 | 0.4<br>(0.3, 0.5) | 1.7<br>(1.2, 2.2) |
|  | 0-9 years | 21,849,433 | 24,763 | 113.3 | 1,087 | 5.0<br>(4.7, 5.3) | 4.4<br>(4.1, 4.6) | 302 | 1.4<br>(1.2, 1.5) | 1.2<br>(1.1, 1.4) |
|  | 10-14 years | 11,112,719 | 3,244 | 29.2 | 129 | 1.2<br>(1.0, 1.4) | 4.0<br>(3.3, 4.6) | 66 | 0.6<br>(0.5, 0.7) | 2.0<br>(1.5, 2.5) |
|  | 15-19 years | 11,072,648 | 9,272 | 83.7 | 339 | 3.1<br>(2.7, 3.4) | 3.7<br>(3.3, 4.0) | 160 | 1.4<br>(1.2, 1.7) | 1.7<br>(1.5, 2.0) |
|  | 10-19 years | 22,185,367 | 12,516 | 56.4 | 468 | 2.1<br>(1.9, 2.3) | 3.7<br>(3.4, 4.1) | 226 | 1.0<br>(0.9, 1.2) | 1.8<br>(1.6, 2.0) |
| **Jan 1<sup>st</sup>-July 10th 2021** |  |  |  |  |  |  |  |  |  |  |
|  | <1 years | 2,119,036 | 6,882 | 324.8 | 141 | 6.7<br>(5.6, 7.8) | 2.0<br>(1.7, 2.4) | 43 | 2.0<br>(1.4, 2.6) | 0.62<br>(0.58, 0.67) |
|  | 1-4 years | 8,613,161 | 1,885 | 21.9 | 35 | 0.4<br>(0.3, 0.5) | 1.9<br>(1.2, 2.5) | 55 | 0.6<br>(0.5, 0.8) | 2.92<br>(2.83, 3.0) |
|  | 5-9 years | 11,000,967 | 1,144 | 10.4 | 34 | 0.3<br>(0.2, 0.4) | 3.0<br>(2.0, 4.0) | 26 | 0.2<br>(0.1, 0.3) | 2.27<br>(2.19, 2.36) |
|  | 0-9 years | 21,733,164 | 9,911 | 45.6 | 210 | 1.0<br>(0.8, 1.1) | 2.1<br>(1.8, 2.4) | 124 | 0.6<br>(0.5, 0.7) | 1.25<br>(1.24, 1.27) |
|  | 10-14 years | 11,083,131 | 1,547 | 14.0 | 44 | 0.4<br>(0.3, 0.5) | 2.8<br>(2.0, 3.7) | 21 | 0.2<br>(0.1, 0.3) | 1.36<br>(1.30, 1.41) |
|  | 15-19 years | 11,076,419 | 4,493 | 40.6 | 174 | 1.6<br>(1.3, 1.8) | 3.9<br>(3.3, 4.4) | 22 | 0.2<br>(0.1, 0.3) | 0.49<br>(0.47, 0.51) |
|  | 10-19 years | 22,159,550 | 6,040 | 27.3 | 218 | 1.0<br>(0.9, 1.1) | 3.6<br>(3.1, 4.1) | 43 | 0.2<br>(0.1, 0.3) | 0.71<br>(0.70, 0.73) |

<sup>a</sup>Data for all-cause deaths come from the excess mortality dataset in Mexico. In these data, COVID-19 deaths were identified if COVID-19 or terms associated with this diagnosis were mentioned on the death certificate. <sup>b</sup>Data come from SISVER with laboratory-confirmed COVID-19 (RT-PCR or antigen).

